# Clinical Characteristics Associated with Functional Seizures in Individuals with Psychosis

**DOI:** 10.1101/2024.10.30.24316444

**Authors:** Allison M. Lake, India A. Reddy, Robert Havranek, Lea K. Davis, Jonah Fox

**Affiliations:** Vanderbilt Genetics Institute, Vanderbilt University Medical Center, Nashville, Tennessee, USA; Division of Genetic Medicine, Department of Medicine, Vanderbilt University Medical Center, Nashville, Tennessee, USA; Department of Psychiatry & Behavioral Sciences, Vanderbilt University Medical Center, Nashville, Tennessee, USA; Department of Neurology, Vanderbilt University Medical Center, Nashville, Tennessee, USA; Division of Data-Driven and Digital Medicine, Department of Medicine, Icahn School of Medicine at Mount Sinai, New York, NY, USA

**Author notes:** **Address for Correspondence:** Allison M. Lake, BSc, Vanderbilt Genetics Institute, Vanderbilt University Medical Center, 2525 West End Avenue, Nashville, TN 37232.

**Keywords:** Functional seizures, psychosis, comorbidities, healthcare utilization

## Abstract

**Background and Hypothesis:** Functional seizures (FS) are episodes characterized by seizure-like events that are not caused by hypersynchronous neuronal activity. Prior studies have suggested an increased prevalence of psychotic disorders among patients with FS, but results have been inconsistent. We hypothesize that FS are associated with psychosis and that among patients with psychosis, the presence of FS may influence patient clinical characteristics, mortality, and medical resource utilization.

**Study Design:** The association between FS and psychosis was assessed using electronic health records data from a total of 752,883 individuals receiving care at Vanderbilt University Medical Center between 1989 and 2023. Analyses of the association between FS and psychiatric outcomes, sexual trauma, healthcare utilization, and other clinical comorbidities were conducted in a subset of 5,239 patients with psychosis.

**Study Results:** Odds of FS were elevated among patients with psychosis compared to controls (OR=10.17, 95% CI=8.55-12.08, p<0.001). Among patients with psychosis, those with FS exhibited higher rates of suicidality (OR=1.98, 95% CI=1.40-2.8, p<0.001), catatonia (OR=1.95, 95% CI=1.23-3.09, p=0.03), sexual trauma history (OR=2.98, 95% CI=2.08-4.26, p<0.001) and had a greater numbers of antipsychotic trials (4.56 versus 3.37, beta=1.16, SE=0.16, p<0.001) than those without FS. Furthermore, patients with comorbid FS had a greater numbers of hospital presentations at one, three, five, and ten years after receiving a psychosis diagnosis (p<0.001).

**Conclusions:** FS are more common among patients with psychosis and are associated with increased healthcare utilization as well as an increased prevalence of suicidality, catatonia, and certain psychiatric and medical comorbidities.

## 1. Introduction

Functional seizures (FS) are paroxysmal events of involuntary altered behavior, awareness and/or responsiveness that resemble epileptic seizures (ES) but are not caused by hypersynchronous neuronal activity.^1^ The DSM-5 classifies FS as a form of functional neurological disorder (FND) or conversion disorder.^2^ Patients with FS have an elevated tendency for somatization and dissociation, as well as pathological emotional regulation, attention, and arousal which may be involved in the pathogenesis of FS.^3^ While several explanatory models have been proposed, the etiology of FS remains relatively poorly understood.^3^ FS is more common in women, and the average age of onset is in the second and third decades of life.^4^ The gold standard for diagnosing FS and distinguishing it from ES is by recording habitual events with video electroencephalography (EEG) monitoring in an epilepsy monitoring unit.^5^ Misdiagnosis is common, and the delay from symptom onset to definitive diagnosis ranges between 3 and 8.4 years.^4^ FS does not respond to medications that are used to treat epilepsy but can improve with different kinds of psychotherapy.^6^ FS is associated with significant direct and indirect healthcare costs including frequent emergency department presentations, hospitalizations, and loss of employment.^7^ Patients with FS have a heightened risk of early death similar to that of patients with ES, which at least in part appears to be explained by the associated comorbidities.^8, 9^

Post-traumatic stress disorder (PTSD), depression, anxiety, chronic pain, insomnia, migraine, and asthma are among the more commonly recognized comorbidities in patients with FS.^1^ Other comorbidities such as psychotic disorders and opioid use were found to explain the increased morality risk found among patients with FS.^9^ A few prior studies suggested that psychotic disorders have an elevated prevalence among patients with FS, but the findings were inconsistent. For instance, a meta-analysis found that the prevalence of psychosis ranged from 0-15% among those with FS.^10^ It is unclear whether the prevalence of FS is elevated among patients with psychosis, but there are reasons to suspect that it may be. For instance, schizophrenia patients demonstrate higher rates of dissociative and functional symptoms when compared to healthy controls.^11, 12^ In addition, traumatic experiences, which are strongly associated with FS, are significantly more common among patients with psychosis compared to the general population.^1, 13, 14^ A systematic review reported that 78.9% of studies found a prevalence of PTSD exceeding 10% among patients with schizophrenia, which is significantly higher than the general population.^15^ There is also some evidence to suggest that patients with psychosis and adverse childhood experiences have worse outcomes including poorer health and increased healthcare resource utilization. ^16–18^ Therefore, we hypothesize that the prevalence of FS is elevated among patients with psychosis and that FS may also impact comorbidities, healthcare resource utilization, psychiatric outcomes, treatment, and mortality in this patient population.

## 2. Methods

### 2.1 Phenotype definitions and validation

Clinical data including demographics, diagnosis and procedure codes, medication records, and clinical notes were extracted from the Synthetic Derivative, a de-identified copy of the Vanderbilt University Medical Center (VUMC) electronic medical record. To mitigate nonrandom missingness between cases and controls, a data floor requiring at least five ICD-9 or ICD-10 codes of any type documented over the age of 18 on separate days across a period at least three years was applied. The psychosis phenotype was defined using a curated list of ICD-9 and ICD-10 codes relating to psychosis (**eTable 1**). A patient was designated as a psychosis “case” if they had documentation, at age 18 or older, of a psychosis diagnosis code on ≥3 separate calendar months or ≥2 separate calendar months plus at least one antipsychotic medication record. Patients with insufficient evidence for a psychosis diagnosis (e.g., only one diagnosis code) or meeting case criteria only before the age of 18 were excluded from the final cohort and all downstream analyses.

The psychosis phenotype definition was validated by manual review of 50 randomly selected patients meeting phenotype criteria. Each patient chart was independently reviewed by a psychiatrist (IAR) and a neurologist (JF) and labeled as having a specific psychotic disorder (schizophrenia; schizoaffective disorder, bipolar type; schizoaffective disorder, depressive type; schizoaffective disorder, unspecified type; bipolar affective disorder with psychotic features, major depressive disorder with psychotic features; delusional disorder; psychosis in other primary psychiatric conditions), having a possible psychotic disorder, or not having a psychotic disorder. Discrepancies between reviewers were resolved through discussion and consensus. Of the 50 patient charts reviewed, 37 (74%) were labeled as having probable psychosis, while 42 (84%) were determined to have a probable or possible psychosis diagnosis.

The functional seizure phenotype was defined using a previously validated algorithm.^19^ Briefly, after excluding patients with specific epilepsy diagnosis codes (for generalized and focal epilepsy) from the analysis, cases were defined as having a diagnosis code for convulsions or conversion disorder, the presence of an FS keyword and the keyword “EEG” in at least one clinical note, and the presence of an EEG procedure code, all documented at age 18 or older. Patients with some evidence of FS but not enough evidence to be classified as a case (e.g., presence of a convulsions code but not an FS keyword; see Goleva et al.) or meeting case criteria only before the age of 18, were excluded from the final cohort and all downstream analyses.^19^ Disclosures of sexual trauma were extracted from clinical notes using matches to key words and phrases from a phenotyping algorithm previously validated at VUMC (algorithm version 2, available at https://phekb.org/phenotype/sexual-assault-disclosures-clinical-notes-v2).^20^

Additional psychiatric outcomes including diagnoses of catatonia or suicidal behavior, electroconvulsive therapy (ECT) procedures (CPT code 90870), and antipsychotic medication trials were extracted using ICD-9 and ICD-10 codes, procedure codes, and medication records, respectively. A full list of codes used can be found in **eTable 1**. For the phenome-wide association analysis (PheWAS), related diagnostic codes were grouped into “phecodes” using the R PheWAS package, requiring two component codes on distinct dates for each diagnosis.^21–23^

Information on inpatient and emergency department (ED) encounters on or after the earliest psychosis diagnosis was extracted for the psychosis cohort healthcare utilization analysis using a combination of visit records and procedure codes. To avoid double-counting encounters, inpatient encounters occurring within three days of one another were combined into a single encounter, and ED encounters within one day of one another were combined into a single encounter. The total number of ED and inpatient encounters for each patient in the psychosis cohort were counted at one, three, five, and ten years from the earliest psychosis diagnosis.

A subset of 100 patients with psychosis and at least one encounter record within one year of the earliest psychosis diagnosis (50 with comorbid FS, 50 with no FS) were randomly sampled, and clinical notes from all encounters within the first year of the earliest psychosis diagnosis were extracted. Of these, clinical notes were only available for 92 patients (47 with comorbid FS, 45 with no FS). Notes were manually reviewed by neurologists JF and RH and medical student AML. Based on clinical note documentation, the diagnostic category of the presenting problem for each encounter was determined. For the purposes of this chart review, a patient was considered to have definite FS if their habitual events were captured by video-EEG recording. If the clinical description of patient events were suspicious for FS and there was no evidence of epileptiform abnormalities on EEG the patient was considered to have suspected FS. This research study was reviewed and approved by the VUMC IRB (IRB#160650) and received a “non-human subjects” determination due to the use of deidentified medical record data.

### 2.2 Statistical analysis

Statistical analyses were conducted in R version 4.2.1.^24^ All regression models included covariates for EHR-recorded sex and either age at earliest psychosis diagnosis (temporally restricted psychosis cohort analyses) or median age at visit (non-temporally restricted analyses). The association between FS and psychosis was tested using logistic regression in the full sample (N=752,883). Further analyses were conducted in the psychosis patient cohort (N=5,239). Differences between psychosis patient groups in outcomes at any EHR time point were assessed using logistic regression for dichotomous outcomes (clozapine and ECT treatment, catatonia diagnoses, codes for suicidal ideation or attempt) or linear regression for continuous outcomes (number of unique antipsychotic medications trialed). PheWAS was performed on 580 phecodes for which there were at least 50 cases in the psychosis sample and at least 5 cases each in males and females. Differences in numbers of encounters at one, three, five, and ten years after the earliest psychosis diagnosis were assessed using Poisson regression with covariates for sex and age at psychosis diagnosis. In the encounter chart review analysis, differences in presenting problems between groups were assessed in separate logistic regressions for each diagnosis category. Time-to-event analysis was performed using Cox proportional hazards regression, and survival curves were generated using the *survival* and *survminer* R packages, respectively.^25–27^

## 3. Results

Analyses were conducted using EHR data from a total of 752,883 individuals receiving care at VUMC between 1989 and 2023, among whom 5,239 individuals with a diagnosis of psychosis and 2,122 with FS were identified (**Table 1**). FS were more common among those with a psychosis diagnosis (2.7%) than those without (0.3%, **Table 1**). Psychosis patients had over ten times the odds of an FS diagnosis after adjusting for sex and age (OR=10.17, 95% CI=8.55-12.08, **Table 2**).

**Table 1.**
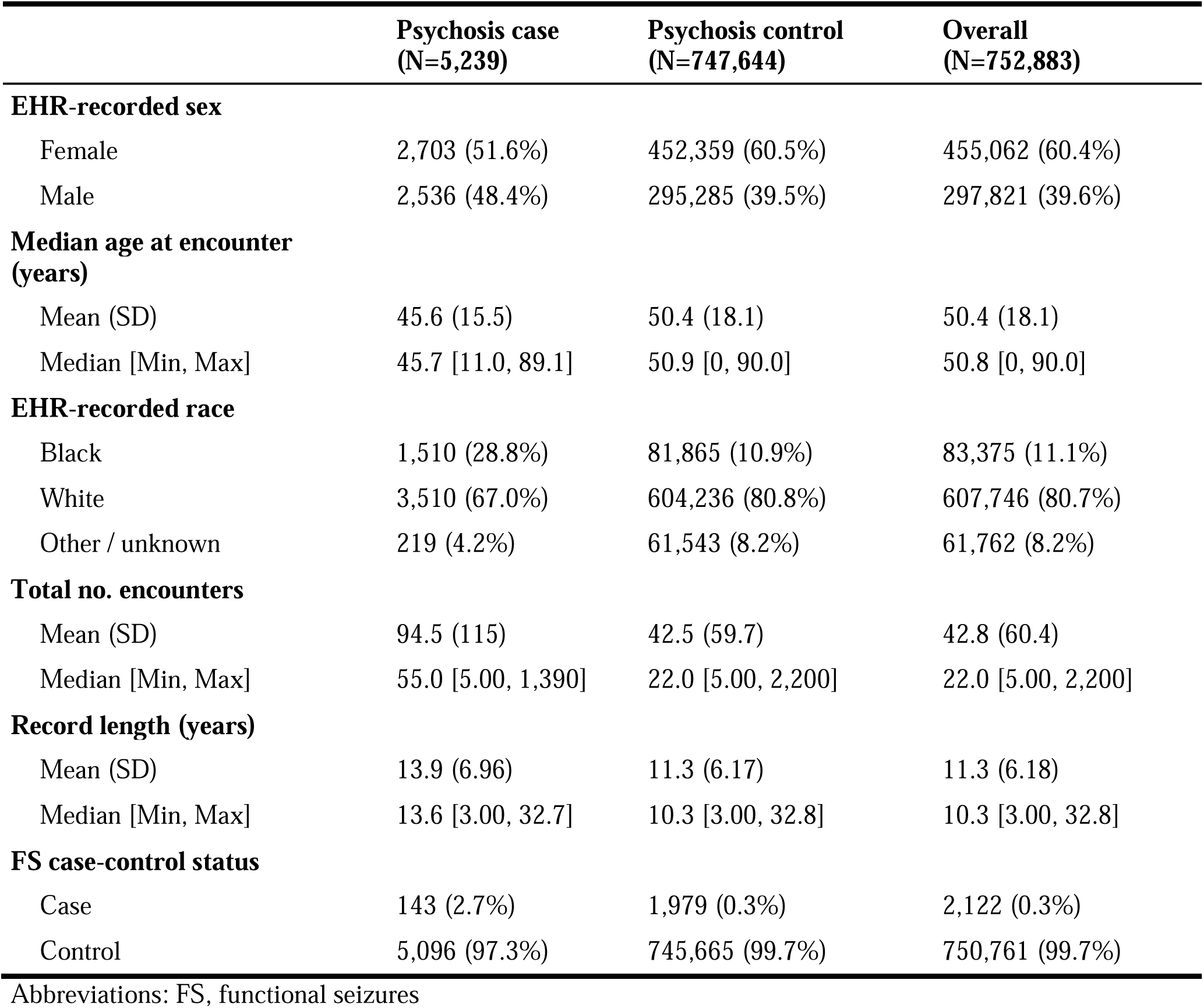
Sample characteristics.

**Table 2.**
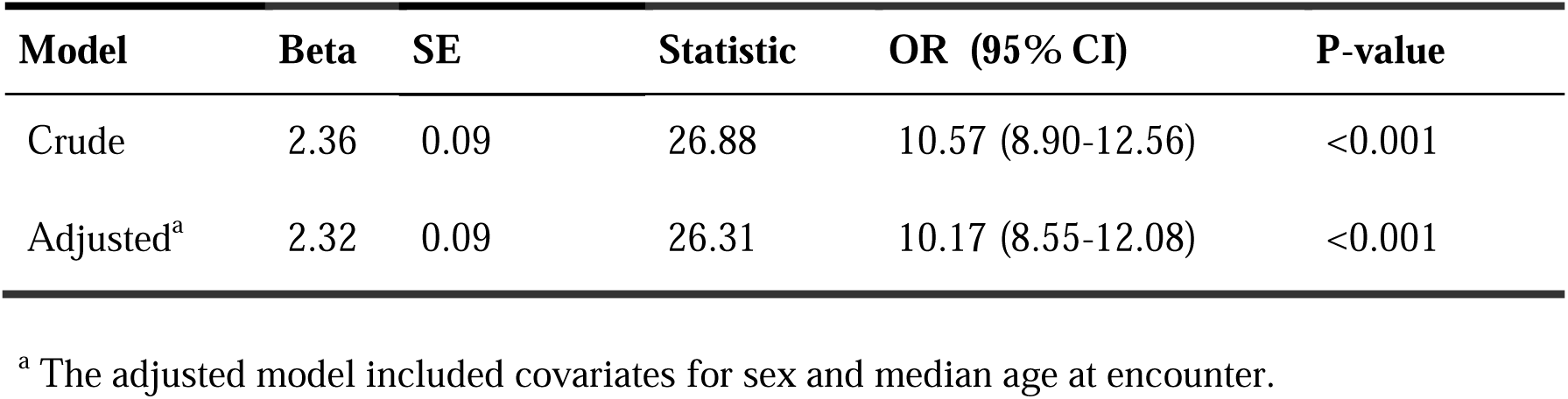
Associations between functional seizures and psychosis in the full EHR sample.

Among the 5,239 patients with a psychosis diagnosis, 67.8% of those with comorbid FS (N=143) were female, as compared with 51.1% of those without FS (χ^2^ (1 d.f., N = 5239) = 14.86, p<0.001, **Table 3**). Psychosis patients with comorbid FS were more likely to receive a diagnostic code for suicidal ideation, suicide attempt, or self-harm (OR=1.98, 95% CI=1.40-2.8, adjusted p<0.001) and had a greater total number of antipsychotic trials on average (4.56 versus 3.37, beta=1.16, SE=0.16, adjusted p<0.001, **Table 4**). Patients with comorbid FS also had a significantly greater odds of having a catatonia diagnosis (OR=1.95, 95 CI=1.23-3.09, adjusted p=0.03) and a history of sexual trauma (OR=2.98, 95% CI=2.08-4.26, p<0.001, **Table 4**) than those without FS. No differences in odds of receiving ECT or clozapine treatment were observed between patient groups.

**Table 3.**
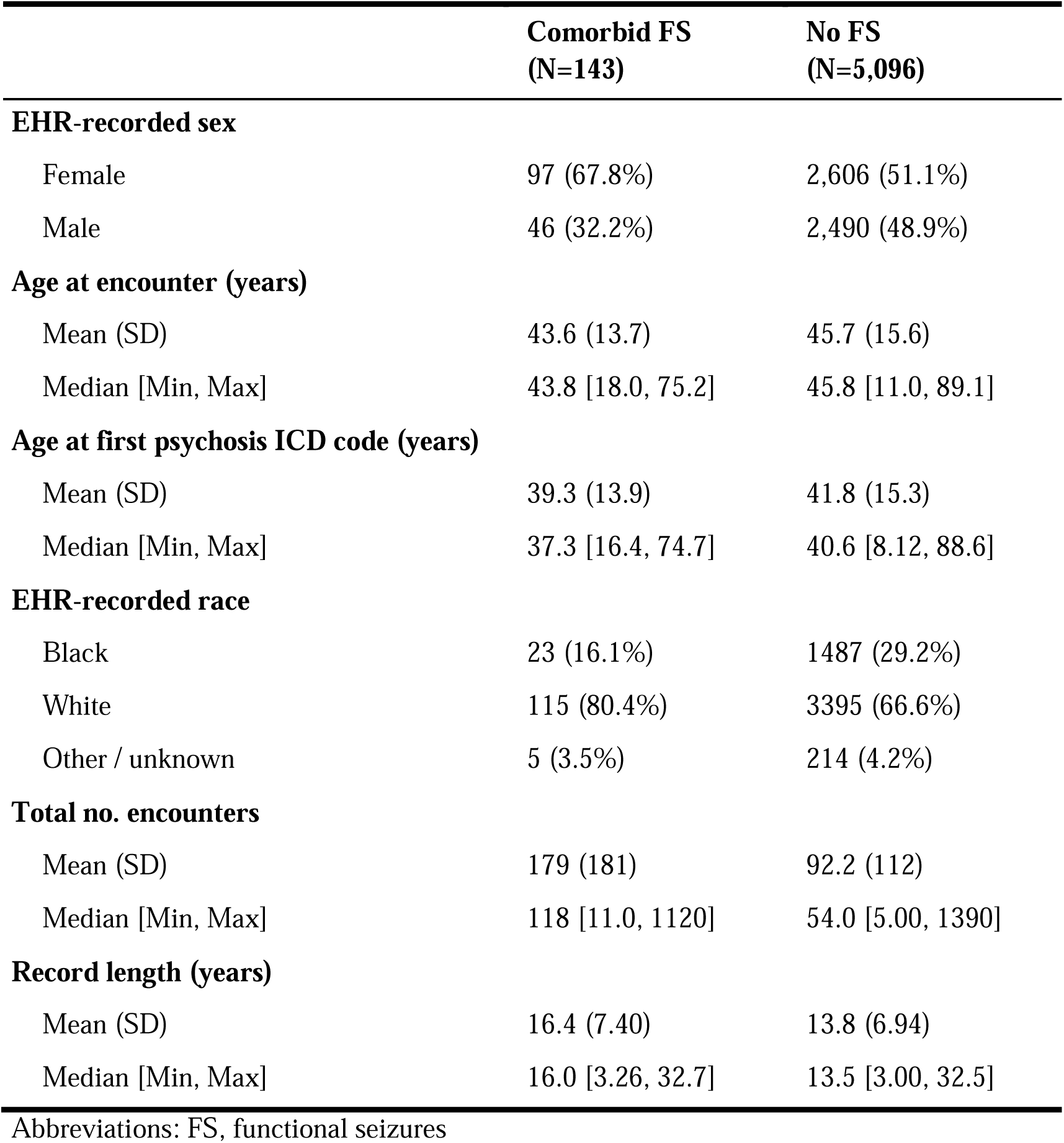
Psychosis cohort sample characteristics.

**Table 4.**
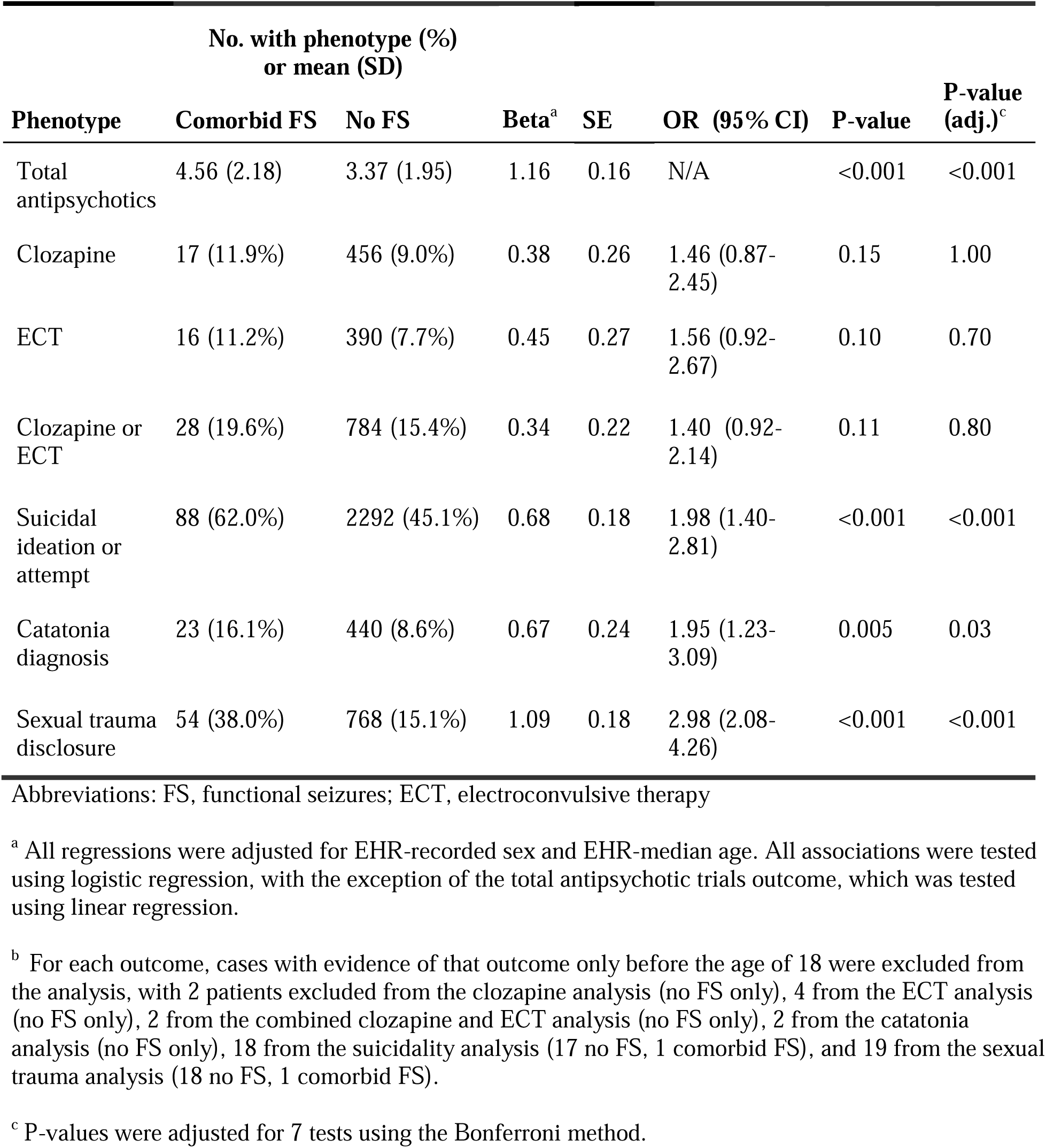
Associations between functional seizures and psychiatric outcomes or phenotypes among patients with psychosis.

Differences in comorbidities between patient groups were assessed using PheWAS. A total of 167 phenotypes were significantly associated with FS among patients with psychosis (p<8.22e-05, corrected for 580 phenotypes tested). Diagnostic categories with the largest number of associated phenotypes included circulatory, neurological, psychiatric, endocrine, and respiratory conditions, as well as injuries and poisonings (**Figure 1**).

**Figure 1.**
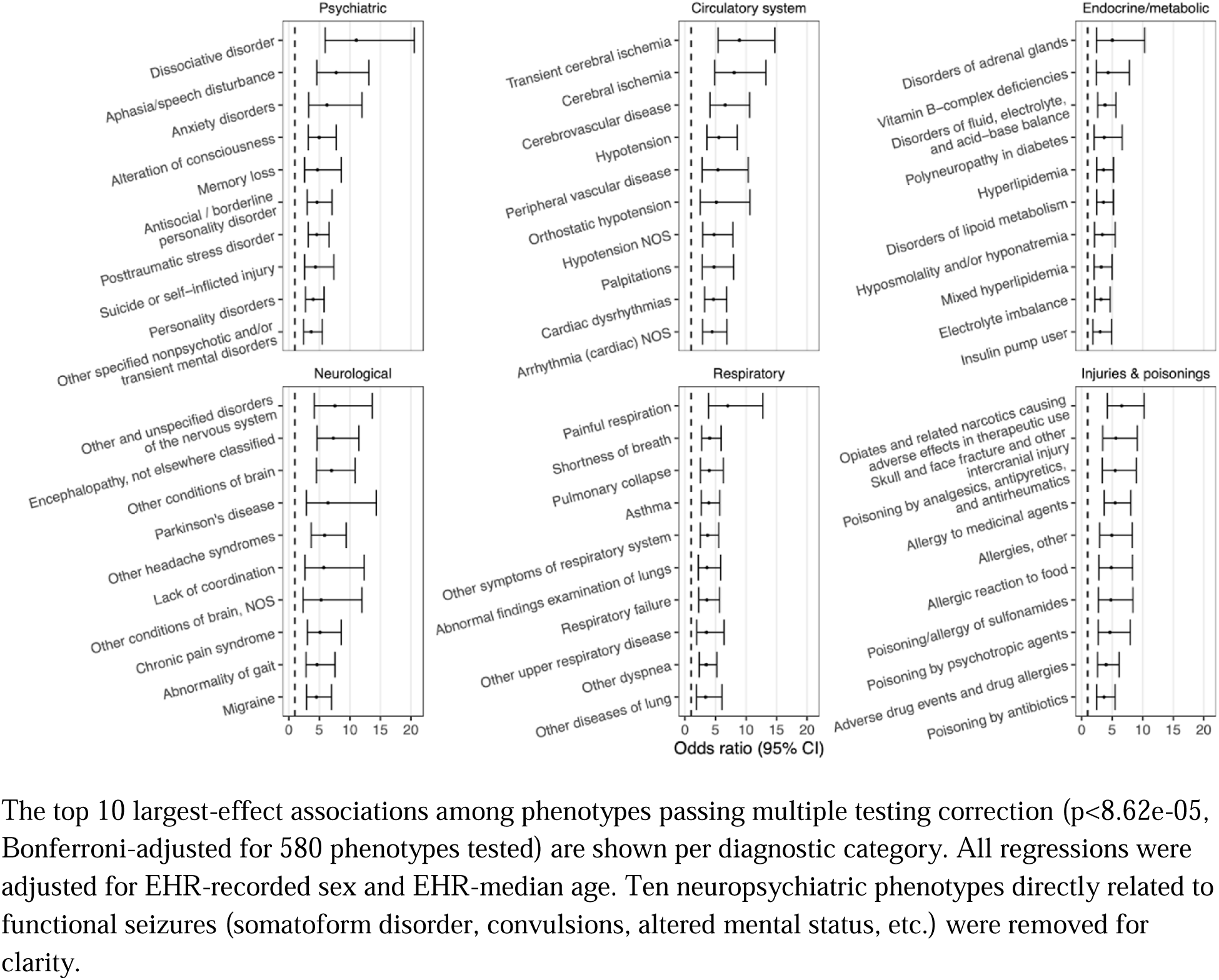
Top ten clinical comorbidities per broad diagnostic category associated with functional seizures among patients with a psychosis diagnosis.

We examined differences in numbers of emergency department (ED) and inpatient encounters between patient groups at time intervals relative to the index psychosis diagnosis. Patients with comorbid FS had significantly greater numbers of inpatient and ED encounters at one, three, five, and ten years after the index psychosis diagnosis (p<0.001, corrected for eight tests using the Bonferroni method, **Table 5**).

**Table 5.**
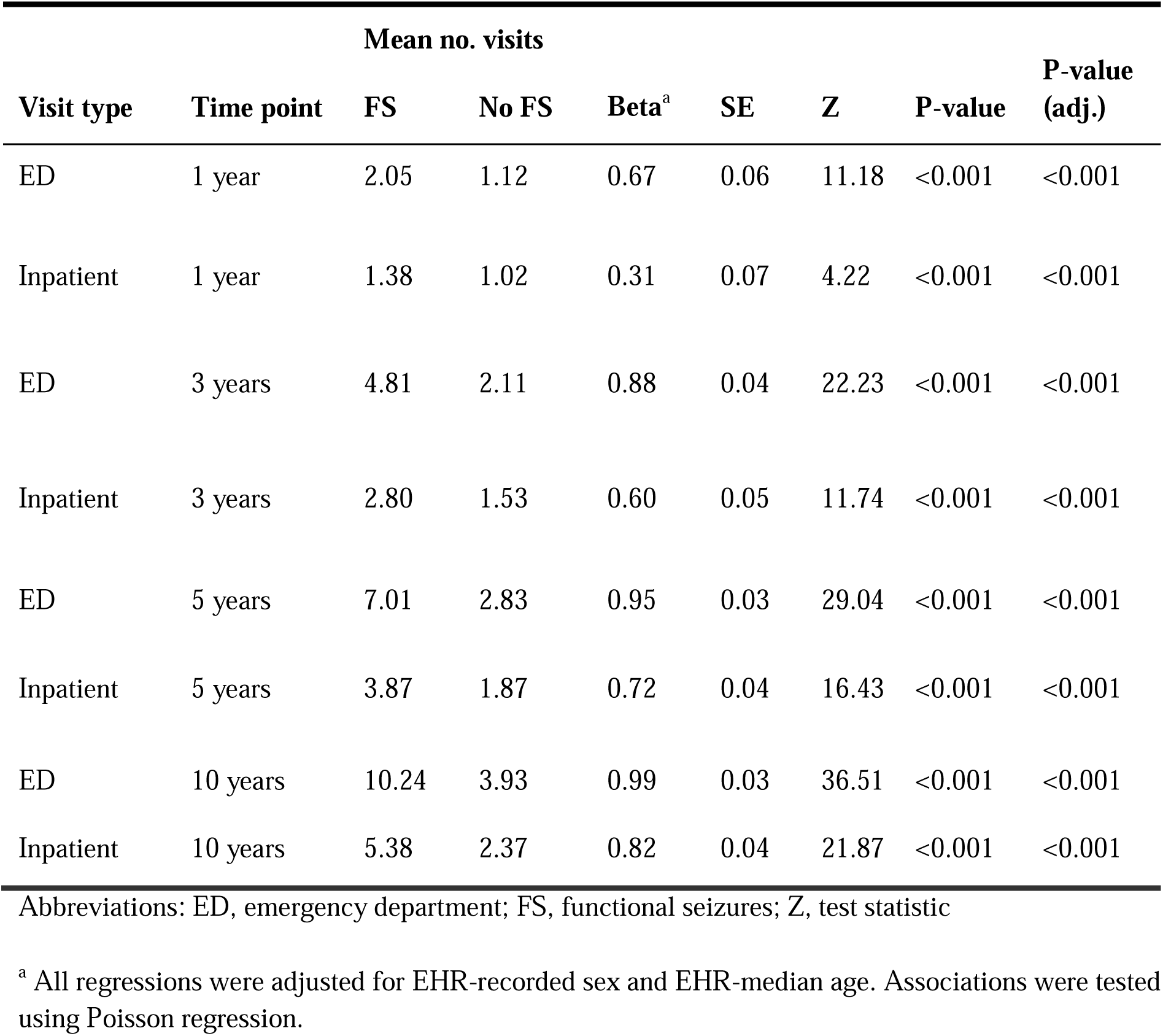
Differences in number of emergency department and inpatient encounters between FS groups, among patients with psychosis.

Clinical notes from a subset of 92 randomly selected individuals (47 with comorbid FS, 45 with psychosis only) were reviewed, and the primary reasons for ED presentations or inpatient hospitalizations within the first year of receiving a psychosis diagnosis were assessed. Across patient groups, the most common reasons for presentation were psychiatric symptoms, injuries and poisonings, circulatory problems, musculoskeletal problems, neurological symptoms, and digestive problems. No differences in the odds of any diagnostic category or of suicidal ideation or attempt were observed between patient groups (**eTable 2, eTable 3**). Of the 47 patients with comorbid FS, 21.3% were determined to have likely or definite functional seizure symptoms as part of their clinical presentation (**eTable 3**).

Survival analysis indicated no significant differences between patient groups in mortality after the earliest psychosis diagnosis in the full sample (hazard ratio = 1.41, 95% CI=0.92-2.15, p=0.12, **eFigure 1**). Given higher mortality rates observed in men, we repeated the analysis stratified by sex and found no significant difference in either group (females, hazard ratio = 1.28, 95% CI=0.75-2.19, p=0.37; males, hazard ratio = 1.69, 95% CI=0.84-3.42, p=0.14, **eFigure 2**). Given that 44 of the 143 individuals (31%) with comorbid FS were first documented to have FS five years or more after the date of their first psychosis diagnosis, we conducted a sensitivity analysis in the sex-combined sample removing these individuals and found similar results (hazard ratio = 1.45, 95% CI=0.87-2.42, p=0.16).

## 4. Discussion

The results of our study suggest that FS is not only more common among patients with psychosis but that its presence may have significant clinical and medical resource utilization implications. Comorbid FS was associated with an increase in inpatient and emergency department presentations, greater numbers of documented antipsychotic trials, catatonia diagnoses, and suicidal ideation or attempts. Furthermore, psychosis patients with comorbid FS were more likely to have a history of sexual trauma than those without FS. Finally, psychosis patients with comorbid FS had significantly different comorbidity profiles.

Psychosis is associated with substantial medical expenditures and healthcare utilization. In the United States, the annual cost attributable to schizophrenia was estimated to be 155.7 billion dollars.^28^ Schizophrenia is a predictor of increased emergency department utilization for both psychiatric and medical disorders and increases likelihood of admission.^29^ FS is also associated with considerable medical resource utilization including frequent emergency department presentations. Our data suggests that the presence of both disorders increases emergency department presentations, hospital admissions, and number of antipsychotic trials compared to psychosis alone.

In a significant proportion of the manually reviewed hospital presentations, the reasons for which psychosis patients—both with and without comorbid FS—presented to the hospital were not directly related to either diagnosis but to other medical or psychiatric indications. Prior research has indicated that a significant proportion of the increased healthcare utilization among both FS and psychosis patients are for the treatment of comorbid conditions.^29–31^ Poor adherence to treatment and inadequate access to outpatient care may account for why some patients may utilize the emergency department,^32^ while adequate treatment of psychosis may potentially reduce healthcare utilization.^33^ Similarly, healthcare utilization significantly declines following both the diagnosis and treatment of FS.^31, 34^ This is notable given that patients with FS often wait years from the time of symptom onset to receiving an accurate diagnosis.^4^ However, we did not evaluate the relationship between healthcare utilization with the timing of receiving a diagnosis of FS or its treatment in this study.

Adverse childhood experiences which may include physical and sexual abuse have been associated with an increased risk for developing psychosis, an earlier onset of schizophrenia, suicidal behavior, and FS.^1, 19, 35^ We found that psychosis patients with comorbid FS were more likely to have a history of sexual trauma than those without FS. This shared risk factor may partially explain the associations between FS, psychosis, and suicidality.^36–38^ Psychosis is well established as a significant risk factor for suicide,^39^ and our results suggest that FS may be associated with a further elevated risk for suicidal ideation or attempts.

Comorbid FS was associated with additional antipsychotic medication trials but no other markers indicative of psychosis severity or treatment responsiveness such as clozapine prescriptions or ECT treatment. Patients with FS may have had more medication trials because they have an increased tendency to report medication allergies, and those with somatization are more likely to report medication intolerances.^40, 41^ Catatonia diagnoses were significantly more common in patients with comorbid FS. Prior work has found that there may be an association between psychological trauma and catatonia which may account for this finding.^42, 43^

There was no significant difference in the primary reason for emergency department presentations and inpatient admissions between the groups among the chart reviewed subset, but the two patient groups did exhibit significantly different medical comorbidity profiles. For instance, patients with comorbid FS were more likely to also have migraine, chronic pain, certain types of respiratory problems such as asthma, allergies, hyperlipidemia, and vascular disease. These conditions have previously been associated with FS.^1, 40, 44, 45^ Psychosis has also been linked to higher prevalences of vascular risk factors and disease.^46, 47^ Chronic pain has a similar prevalence in patients with schizophrenia and the general population, but it is speculated that it may be underdiagnosed due to differences in pain perception and under-reporting.^48^

Both psychosis and FS are associated with an elevated risk of mortality.^8, 9, 49^ Our results suggested that the mortality risk of psychosis patients was not significantly different based on the presence or absence of comorbid FS. Given that the elevated mortality risk in FS appears to be driven by associated comorbidities and that psychosis is also associated with mortality there may be no significant added risk when both conditions are present.^9^ On the other hand, it is possible that we had an insufficient sample size to detect a difference and/or that there is a difference in certain subgroups. We examined survival curves stratified by sex and found that neither male nor female psychosis patients with FS had an increased mortality risk.

Limitations of this study include the use of diagnosis codes to identify patients and the retrospective design of the study. However, the algorithms used to identify patients with psychosis and FS were validated and found to have an adequate positive predictive value. The sample size of patients with psychosis and comorbid FS was relatively small, which may have limited our sensitivity to detect certain findings. Additionally, our mortality analysis was limited by the information available in the electronic medical record, and it is probable that there was missing data.

## 5. Conclusion

Psychosis patients have an increased prevalence of FS, which is associated with unique clinical characteristics including an elevated use of medical resources, suicidality, and certain comorbidities. Increased awareness and screening of psychosis patients for comorbid FS may assist in the facilitation of optimal care. Future research could evaluate how treatment of either disorder affects outcomes in this unique group of patients.

## Supporting information

eTable 1

## Data Availability

Data are available from Vanderbilt University Medical Center with institutional restrictions that apply to the acquisition, use, and dissemination of data. To request reasonable access to data for work conducted in a non-profit academic setting, please contact the Vanderbilt Institute for Clinical and Translational Research and request an application to the Integrated Data Access and Services Core.

## References

1. Popkirov S, Asadi Pooya AA, Duncan R, et al. The aetiology of psychogenic non epileptic seizures: risk factors and comorbidities. Epileptic disorders 2019;21(6):529–547.

2. American Psychiatric Association D, American Psychiatric Association D. Diagnostic and statistical manual of mental disorders: DSM-5. Vol 5: American psychiatric association Washington, DC; 2013.

3. Brown RJ, Reuber M. Towards an integrative theory of psychogenic non-epileptic seizures (PNES). Clinical psychology review 2016;47:55–70.

4. Bompaire F, Barthelemy S, Monin J, Quirins M, Marion L, Smith C, Boulogne S, Auxemery Y. PNES epidemiology: what is known, what is new? European Journal of Trauma & Dissociation 2021;5(1):100136.

5. LaFrance Jr WC, Baker GA, Duncan R, Goldstein LH, Reuber M. Minimum requirements for the diagnosis of psychogenic nonepileptic seizures: a staged approach: a report from the International League Against Epilepsy Nonepileptic Seizures Task Force. Epilepsia 2013;54(11):2005–2018.

6. Lopez MR, LaFrance WC. Treatment of psychogenic nonepileptic seizures. Current neurology and neuroscience reports 2022;22(8):467–474.

7. Stephen CD, Fung V, Lungu CI, Espay AJ. Assessment of emergency department and inpatient use and costs in adult and pediatric functional neurological disorders. JAMA neurology 2021;78(1):88–101.

8. Nightscales R, McCartney L, Auvrez C, et al. Mortality in patients with psychogenic nonepileptic seizures. Neurology 2020;95(6):e643–e652.

9. Tan M, Pearce N, Tobias A, Cook MJ, D’Souza WJ. Influence of comorbidity on mortality in patients with epilepsy and psychogenic nonepileptic seizures. Epilepsia 2023;64(4):1035–1045.

10. Diprose W, Sundram F, Menkes DB. Psychiatric comorbidity in psychogenic nonepileptic seizures compared with epilepsy. Epilepsy & Behavior 2016;56:123–130.

11. Kanchanatawan B, Sirivichayakul S, Thika S, et al. Physio-somatic symptoms in schizophrenia: association with depression, anxiety, neurocognitive deficits and the tryptophan catabolite pathway. Metabolic brain disease 2017;32:1003–1016.

12. Schäfer I, Fisher HL, Aderhold V, et al. Dissociative symptoms in patients with schizophrenia: relationships with childhood trauma and psychotic symptoms. Comprehensive psychiatry 2012;53(4):364–371.

13. Martin D, Philips M, Greenstone H, Davies J, Stewart G, Ewins E, Zammit S. Examining the Relationship Between Trauma, Post-Traumatic Stress Disorder and Psychosis in Patients in a UK Secondary Care Service. Psychiatr Res Clin Pract Summer 2023;5(2):51–59.

14. Hardy KV, Mueser KT. Editorial: Trauma, Psychosis and Posttraumatic Stress Disorder. Front Psychiatry 2017;8:220.

15. Dallel S, Cancel A, Fakra E. Prevalence of posttraumatic stress disorder in schizophrenia spectrum disorders: a systematic review. Neuropsychiatry 2018;8(3):1027–1037.

16. Rosenberg SD, Lu W, Mueser KT, Jankowski MK, Cournos F. Correlates of adverse childhood events among adults with schizophrenia spectrum disorders. Psychiatric services 2007;58(2):245–253.

17. Trotta A, Murray R, Fisher H. The impact of childhood adversity on the persistence of psychotic symptoms: a systematic review and meta-analysis. Psychological medicine 2015;45(12):2481–2498.

18. Chartier MJ, Walker JR, Naimark B. Separate and cumulative effects of adverse childhood experiences in predicting adult health and health care utilization. Child abuse & neglect 2010;34(6):454–464.

19. Goleva SB, Lake AM, Torstenson ES, Haas KF, Davis LK. Epidemiology of Functional Seizures Among Adults Treated at a University Hospital. JAMA Netw Open Dec 1 2020;3(12):e2027920.

20. Lake AM, Goleva SB, Samuels LR, Carpenter LM, Davis LK. Sex Differences in Health Conditions Associated with Sexual Assault in a Large Hospital Population. Complex Psychiatry Jan 2023;8(3-4):80–89.

21. Denny JC, Ritchie MD, Basford MA, et al. PheWAS: demonstrating the feasibility of a phenome-wide scan to discover gene-disease associations. Bioinformatics May 1 2010;26(9):1205–1210.

22. Carroll RJ, Bastarache L, Denny JC. R PheWAS: data analysis and plotting tools for phenome-wide association studies in the R environment. Bioinformatics Aug 15 2014;30(16):2375–2376.

23. Wu P, Gifford A, Meng X, et al. Mapping ICD-10 and ICD-10-CM Codes to Phecodes: Workflow Development and Initial Evaluation. JMIR Med Inform Nov 29 2019;7(4):e14325.

24. R: A language and environment for statistical computing. Version 4.2.1. Vienna, Austria; 2022.

25. Therneau T. A Package for Survival Analysis in R. Version: R package version 3.3-1. Available at: https://CRAN.R-project.org/package=survival.

26. Therneau TM, Grambsch PM. Modeling Survival Data: Extending the Cox Model: Springer New York; 2013.

27. *survminer: Drawing Survival Curves using ’ggplot2* [computer program]. Version: R package version 0.4.9; 2021. Available at: https://CRAN.R-project.org/package=survminer.

28. Cloutier M, Aigbogun MS, Guerin A, et al. The Economic Burden of Schizophrenia in the United States in 2013. J Clin Psychiatry Jun 2016;77(6):764–771.

29. Ronaldson A, Elton L, Jayakumar S, Jieman A, Halvorsrud K, Bhui K. Severe mental illness and health service utilisation for nonpsychiatric medical disorders: a systematic review and meta-analysis. PLoS medicine 2020;17(9):e1003284.

30. Ramamurthy S, Steven Brown L, Agostini M, et al. Emergency department visits and readmissions in patients with psychogenic nonepileptic seizures (PNES) at a safety net hospital. Epilepsy & Behavior 2021/09/01/ 2021;122:108225.

31. Salinsky M, Storzbach D, Goy E, Kellogg M, Boudreau E. Health care utilization following diagnosis of psychogenic nonepileptic seizures. Epilepsy & Behavior 2016/07/01/ 2016;60:107–111.

32. Hardy M, Jackson C, Byrne J. Antipsychotic adherence and emergency department utilization among patients with schizophrenia. Schizophrenia Research 2018;201:347–351.

33. Okoli CTC, Kappi A, Wang T, Makowski A, Cooley AT. The effect of long-acting injectable antipsychotic medications compared with oral antipsychotic medications among people with schizophrenia: A systematic review and meta-analysis. International Journal of Mental Health Nursing 2022;31(3):469–535.

34. Deleuran M, Nørgaard K, Andersen NB, Sabers A. Psychogenic nonepileptic seizures treated with psychotherapy: Long-term outcome on seizures and healthcare utilization. Epilepsy & Behavior 2019/09/01/ 2019;98:195–200.

35. Berardelli I, Rogante E, Sarubbi S, Erbuto D, Lester D, Pompili M. The importance of suicide risk formulation in schizophrenia. Frontiers in psychiatry 2021;12:779684.

36. Yates K, Lång U, Cederlöf M, Boland F, Taylor P, Cannon M, McNicholas F, DeVylder J, Kelleher I. Association of Psychotic Experiences With Subsequent Risk of Suicidal Ideation, Suicide Attempts, and Suicide Deaths: A Systematic Review and Meta-analysis of Longitudinal Population Studies. JAMA Psychiatry Feb 1 2019;76(2):180–189.

37. Chapman CL, Mullin K, Ryan CJ, Kuffel A, Nielssen O, Large MM. Meta-analysis of the association between suicidal ideation and later suicide among patients with either a schizophrenia spectrum psychosis or a mood disorder. Acta Psychiatr Scand Mar 2015;131(3):162–173.

38. Gupta R, Garg D, Kumar N, Singh MB, Shukla G, Goyal V, Pandey RM, Srivastava AK. Psychiatric co-morbidities and factors associated with psychogenic non-epileptic seizures: a case–control study. Seizure 2020/10/01/ 2020;81:325–331.

39. Girgis RR. The neurobiology of suicide in psychosis: A systematic review. Journal of Psychopharmacology 2020;34(8):811–819.

40. Robbins NM, Larimer P, Bourgeois JA, Lowenstein DH. Number of patient-reported allergies helps distinguish epilepsy from psychogenic nonepileptic seizures. Epilepsy & behavior : E&B Feb 2016;55:174–177.

41. Hassel JC, Danner D, Hassel AJ. Psychosomatic or allergic symptoms? High levels for somatization in patients with drug intolerance. J Dermatol Oct 2011;38(10):959–965.

42. Ross CA, Browning E. The relationship between catatonia and dissociation: A preliminary investigation. Journal of Trauma & Dissociation 2016;17(4):426–434.

43. Biles TR, Anem G, Youssef NA. Should catatonia be conceptualized as a pathological response to trauma? The Journal of Nervous and Mental Disease 2021;209(5):320–323.

44. Fox J, Goleva SB, Haas KF, Davis LK. Functional seizures are associated with cerebrovascular disease and functional stroke is more common in patients with functional seizures than epileptic seizures. Epilepsy & behavior : E&B Mar 2022;128:108582.

45. Fox J, Mishra M. Hypertension and other vascular risk factors in patients with functional seizures. Epilepsy & behavior : E&B Mar 2024;152:109650.

46. Li X, Hu S, Liu P. Vascular-related biomarkers in psychosis: a systematic review and meta-analysis. Frontiers in Psychiatry 2023;14:1241422.

47. Galletly CA, Foley DL, Waterreus A, Watts GF, Castle DJ, McGrath JJ, Mackinnon A, Morgan VA. Cardiometabolic risk factors in people with psychotic disorders: the second Australian national survey of psychosis. Australian & New Zealand Journal of Psychiatry 2012;46(8):753–761.

48. Stubbs B, Mitchell AJ, De Hert M, Correll CU, Soundy A, Stroobants M, Vancampfort D. The prevalence and moderators of clinical pain in people with schizophrenia: A systematic review and large scale meta-analysis. Schizophrenia Research 2014/12/01/ 2014;160(1):1–8.

49. Oakley P, Kisely S, Baxter A, Harris M, Desoe J, Dziouba A, Siskind D. Increased mortality among people with schizophrenia and other non-affective psychotic disorders in the community: a systematic review and meta-analysis. Journal of psychiatric research 2018;102:245–253.

