## Supplement for "Clinical Characteristics Associated with Functional Seizures in Individuals with Psychosis"

**eTable 2.** Associations between functional seizures and reasons for emergency department presentations or inpatient hospitalizations in the year after the first psychosis diagnosis (among a chart-reviewed subset)

| **Diagnosis group**^a^ | **No. patients with**  **presenting problem (%)** | | **OR (95% CI)**^b^ | **P-value** |
| --- | --- | --- | --- | --- |
|  | **Comorbid FS**  **N=47** | **No FS**  **N=45** |  |  |
| Circulatory problem  (including stroke) | 5 (10.6%) | 8 (17.8%) | 0.60 (0.17-2.07) | 0.42 |
| Injury and poisoning | 11 (23.4%) | 7 (15.6%) | 1.72 (0.59-5.00) | 0.32 |
| Musculoskeletal problem | 7 (14.9%) | 5 (11.1%) | 1.36 (0.39-4.69) | 0.63 |
| Psychiatric symptom^c^ | 33 (70.2%) | 32 (71.1%) | 0.94 (0.38-2.32) | 0.89 |
| Neurological symptom (excluding stroke) | 8 (17.0%) | 4 (8.9%) | 2.17 (0.59-8.06) | 0.25 |
| Digestive problem | 6 (12.8%) | 5 (11.1%) | 1.15 (0.32-4.14) | 0.83 |

^a^ A patient was included in a diagnostic group if the patient presented with that problem at any point during the one-year period. Some patients presented with different problems at different visits, and thus the diagnostic groups are not mutually exclusive.

^b^ All regressions were adjusted for sex and age at first psychosis diagnosis code.

^c^ Presentations in which definite or suspected functional seizures was the primary complaint were categorized as psychiatric symptoms.

**eTable 3.** Associations between functional seizures and additional features of presentations or inpatient hospitalizations in the year after the first psychosis diagnosis (among a chart-reviewed subset)

| **Diagnosis group**^a^ | **No. patients with**  **presentation feature noted (%)** | | **OR (95% CI)**^a^ | **P-value** |
| --- | --- | --- | --- | --- |
|  | **Comorbid FS**  **N=47** | **No FS**  **N=45** |  |  |
| Suspected or definite functional seizures | 10 (21.3%) | 0 | N/A | N/A |
| Definite functional seizures | 4 (8.5%) | 0 | N/A | N/A |
| Suicidal ideation | 18 (38.3%) | 21 (46.7%) | 0.71 (0.31-1.64) | 0.42 |
| Suicide attempt or self-injurious behavior | 8 (17.0%) | 3 (6.7%) | 2.90 (0.71-11.80) | 0.14 |

^a^ All regressions were adjusted for sex and age at first psychosis diagnosis code.

**eFigure 1.** Survival probability after index psychosis diagnosis by patient group

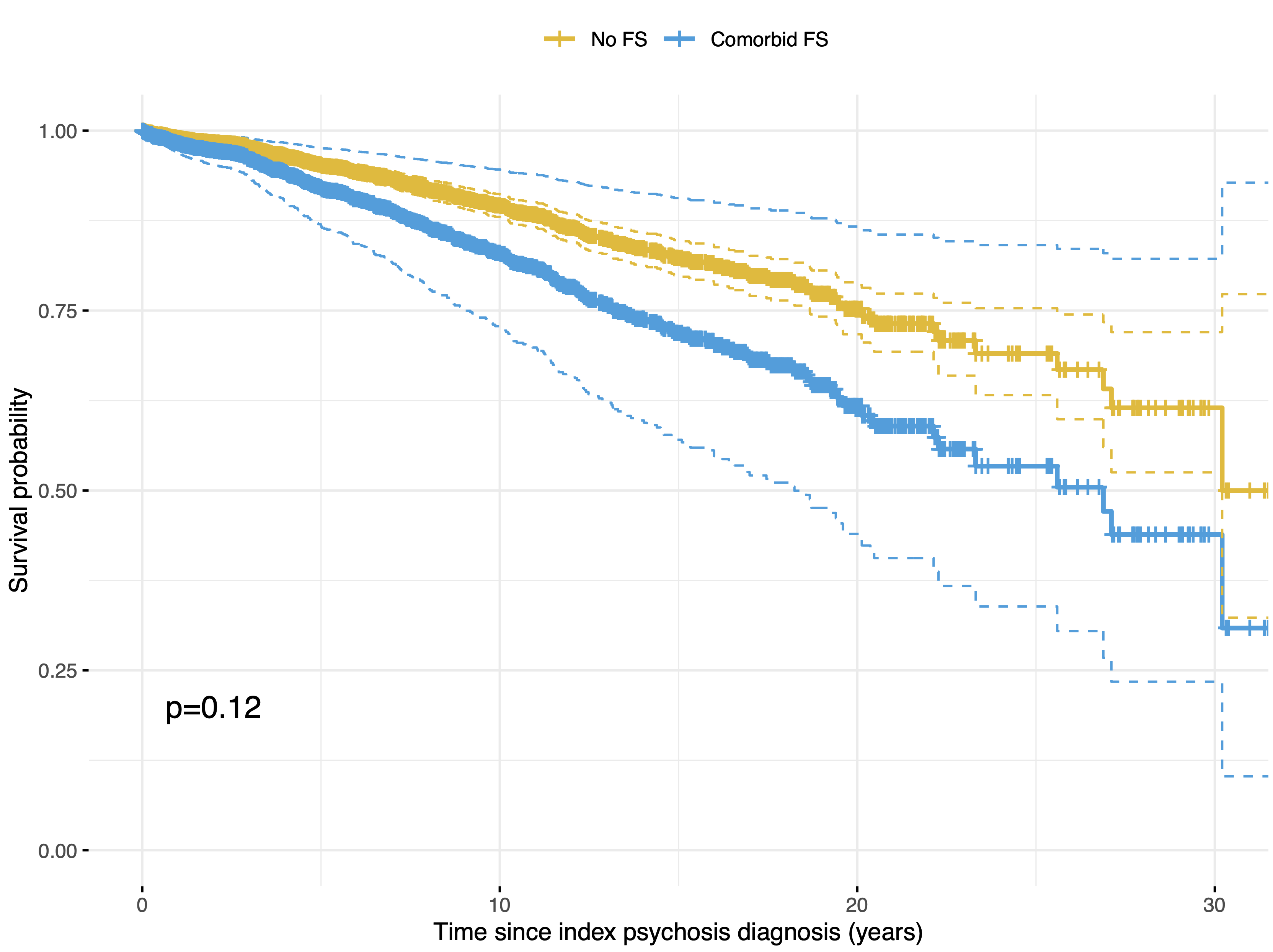

Survival time after the earliest psychosis diagnostic code was assessed in those with comorbid functional seizures and psychosis (comorbid FS) versus those with psychosis only (no FS). Multivariable Cox proportional hazards models were fit with covariates for FS status, age at index diagnosis, and sex. No significant differences in survival time were observed between groups (hazard ratio = 1.41, 95% CI= 0.92-2.15, p=0.12). Predicted survival curves for each patient group were generated using the average age at index diagnosis (41.8 years) and proportion of males (48.4%) as the age and sex covariates, respectively.

**eFigure 2.** Survival probability after index psychosis diagnosis by patient group, stratified by sex

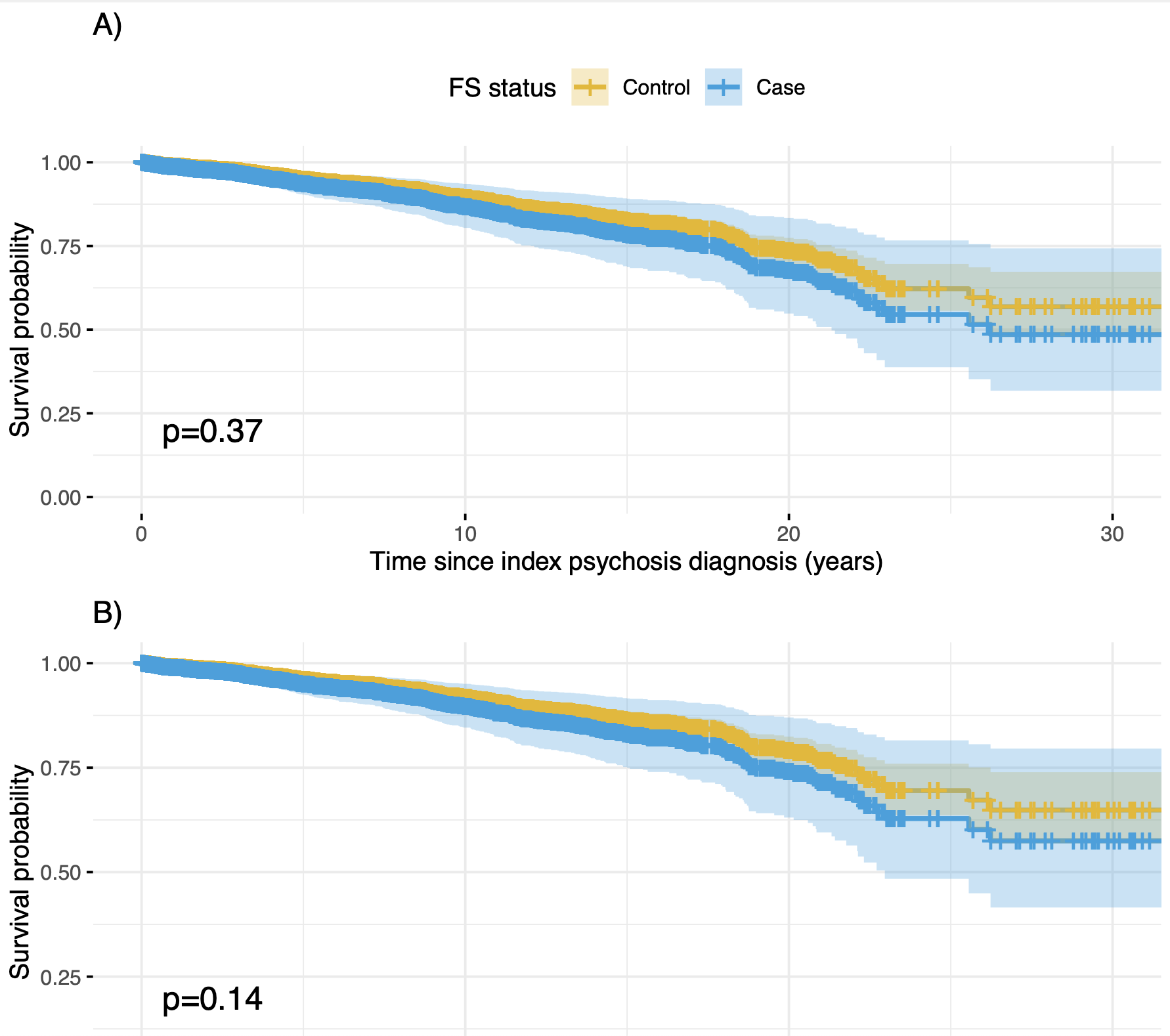

Survival time after the earliest psychosis diagnostic code was assessed in those with comorbid functional seizures and psychosis (comorbid FS) versus those with psychosis only (no FS), separately in females (**A**) and males (**B**). Multivariable Cox proportional hazards models were fit with covariates for FS status and age at index diagnosis. No significant differences in survival time were observed between groups in females (hazard ratio = 1.28, 95% CI=0.75-2.19, p=0.37) or males (hazard ratio = 1.69, 95% CI=0.84-3.42, p=0.14). Predicted survival curves for each patient group were generated using the average age at index diagnosis (43.9 years for females, 39.5 for males) as the age covariates.
